# Short and Long-term Mortality of Pneumococcal Pneumonia in Adults and Children- A Detailed Scoping Review

**DOI:** 10.1101/2023.07.15.23292707

**Authors:** Rupalakshmi Vijayan, Shavy Nagpal, Manthi Dissanayake, Shameera Shaik Masthan, Aiman Fatima, Julio Ramirez, Sahana Karkera, Ipshita Dutta, Pavani Karani, Andrea Galecio Chao

## Abstract

Streptococcus pneumoniae is a common infectious agent responsible for pneumonia, which can result in serious complications such as meningitis, sepsis, morbidity, and mortality. The objective of this study is to examine the risk factors and complications associated with mortality caused by community-acquired pneumococcal pneumonia in both adults and children. To conduct this research, a thorough literature review was carried out by researchers between May 1, 2021, and August 1, 2021. Various databases, including PubMed, WHO, clinicaltrials.gov, Embase, Web of Science, Cochrane, and Google Scholar, were searched using specific keywords such as mortality, pneumococcal pneumonia, adults, children, and their combinations. The study encompassed patients of all age groups affected by pneumococcal pneumonia, while systematic reviews focusing on other types of pneumonia and non-pneumonia patients were excluded. After eliminating duplicate studies, the search yielded 1783 relevant articles, which underwent title and abstract screening. Ultimately, 8 studies were included in the final review. In conclusion, pneumococcal pneumonia is a significant contributor to mortality among both adults and children. This research emphasizes the importance of implementing effective management strategies to reduce long-term mortality

## Introduction

Streptococcus pneumoniae is a bacterial pathogen known to cause pneumonia, which can manifest as invasive pneumococcal disease (IPD) and severe non-invasive lower respiratory disease, both of which can be fatal.[1] Among cases of community-acquired pneumonia (CAP) caused by pneumococcus, approximately 20-30% are classified as invasive pneumococcal pneumonia (invasive pneumonia), while the remaining 70-80% are considered non-invasive pneumonia.[2] Pneumococcal infection is a significant global health concern, contributing to substantial morbidity and mortality.[1] Individuals with chronic conditions such as chronic obstructive pulmonary disease, bronchial asthma, chronic cardiovascular disease, cerebrovascular disease, chronic renal disease, chronic liver disease, or diabetes mellitus, as well as the elderly and immunocompromised, face an increased risk of acquiring pneumococcal pneumonia (PP) and bacteremia.[3] Bacterial pneumococcal pneumonia (BPP) can lead to various complications, ranging from adult respiratory distress syndrome to septic shock and death.[1] Numerous guidelines have been established to promote standardized management of pneumonia, with several studies demonstrating that adherence to these guidelines has resulted in reduced mortality rates.[4]

Over the years, advancements in early detection methods, appropriate antibiotic administration, and consistent clinical management have contributed to a decline in the case fatality rate among patients with pneumococcal pneumonia.[5] This study aims to provide a comprehensive overview of current trends in the prediction and treatment of pneumococcal pneumonia. The research questions guiding this study are (1) to assess short and long-term mortality in adults and children affected by pneumococcal pneumonia and (2) to evaluate the risk factors and complications associated with mortality caused by pneumococcal pneumonia.

## Methods

To gather relevant literature, a search was conducted from January 2016 to May 31st, 2021, utilizing seven databases, namely PubMed, WHO, clinicaltrials.gov, Embase, Web of Science, Cochrane, and Google Scholar. The search terms used included keywords such as Mortality, Pneumococcal Pneumonia, adults, children, and various combinations thereof. The study encompassed patients of all age groups affected by pneumococcal pneumonia, while systematic reviews focusing on other types of pneumonia and non-pneumonia patients were excluded. A comprehensive search strategy, employing an umbrella method, was utilized, and the reference lists of retrieved articles were cross-checked. After the exclusion process, eight articles were selected for the final review. These selected articles were individually reviewed by two authors, and any disagreements were resolved through discussions with a third reviewer, ultimately reaching a consensus among the authors. The review search encompassed studies conducted in different countries worldwide. To eliminate duplicate studies, Endnote X9 software was employed.

## Ethical approval

Ethical approval was not required for this study, as public databases were used for obtaining data and patients were not directly involved.

## Results

In the 8 studies reviewed, involving 19,137 patients (Study 1-8), significant findings were observed:

A retrospective study in Barcelona from 1997-2016 (Study 1) with 6,403 patients found an overall 30-day mortality rate of 8% for pneumococcal pneumonia. There were no significant changes in mortality over different 5-year periods. A prospective study in Brazil from 2009-2017 (Study 2) involving 186 patients compared adults and elderly individuals with pneumococcal pneumonia. The elderly had a higher 30-day mortality rate but fewer ICU admissions and shorter in-patient stays. An epidemiological study in Europe in 2015 (Study 3) included 21,118 confirmed pneumonia cases, reporting a mortality rate of 14%. Patients with chronic respiratory disorders had a higher risk of bacteremic pneumonia, but their mortality risk was not higher than those without this comorbidity.

Chronic lung illness, such as COPD and bronchial asthma, was identified as an independent risk factor for pneumococcal pneumonia, especially in the elderly (Study 4). Chronic renal illness did not significantly affect mortality in relation to pneumococcal pneumonia (Study 5). Smoking was not found to have a significant effect on the occurrence of pneumococcal pneumonia in the studied age groups (Study 6). A study in the Netherlands from 2004-2012 (Study 7) with 228 invasive pneumonia patients revealed that comorbidities such as chronic cardiac, lung, and hepatic disorders, renal failure, immunosuppression, chronic neurological diseases, HIV, diabetes mellitus, tobacco usage, and alcohol addiction were associated with pneumococcal disease-related mortality.

These findings highlight the impact of various risk factors and comorbidities on mortality rates in pneumococcal pneumonia patients (Study 1-7), [6-25].

## Discussion

The most common comorbidities in the studied population were chronic obstructive pulmonary disease (COPD) and bronchial asthma, which were associated with an increased risk of bacteremic pneumonia (adjusted OR: 3.07, 95% CI: 1.23–7.65, p 0.01). Adults with asthma had a 12-17% attributable risk of invasive pneumococcal infections, especially with frequent asthma exacerbations [25,26]. Pneumonia in dialysis patients occurred at a rate of 27.9 per 100 people per year, with a 1-year survival rate of 0.51 [25]. Chronic renal failure was more prevalent in people over 65 years old but did not affect mortality [26]. Splenic volume less than 40 cm3 was found to be significantly associated with mortality within 30 days in patients with pneumococcal pneumonia [30,31,32,33,34]. Alcohol misuse and acid-suppressing drugs were associated with increased mortality [30]. Smoking was not significantly associated with pneumonia in the studied age groups [39,40,41,42]. Childhood pneumococcal vaccination had indirect effects on the incidence of pneumococcal disease [46]. Risk factors for mortality in children with pneumonia included convulsions, age <12 months, lower weight, unsafe water consumption, lower parental education, severity of disease, and presence of co-morbid conditions [47,48,49,50,51].

Greater mortality in children under the age of five was positively associated with the severity of pneumonia [51,52]. Surprisingly, there was no significant link found between pentavalent vaccination status and pneumonia mortality. However, pentavalent vaccination was found to be protective against severe pneumonia. Establishing a link between immunization and mortality was challenging due to the low number of pneumonia deaths and potential misclassification of vaccination status based on parental reports. Modifiable factors such as weight for age and availability of safe drinking water were associated with both severity and death. Additionally, parental education level and the omission of more severe and younger individuals from the study may have contributed to discrepancies. Higher maternal education has been previously associated with improved health outcomes in children, including lower childhood mortality rates [53].

### Limitations

Our research has limitations that should be considered. Firstly, we did not have information on cause-specific mortality. Secondly, the severity of in-hospital issues was not extensively documented, and we only knew whether they occurred. Thirdly, our findings may not be applicable to patients with pneumococcal bacteremia without pneumonia or other invasive pneumococcal diseases like meningitis. Fourthly, our study focused on patients with BPP who were treated before the implementation of conjugated vaccinations in addition to the standard pneumococcal polysaccharide vaccine for this patient population [52,53]. Fifthly, we lacked detailed data on antibiotic administration, including timing, route, dose, frequency, and antimicrobial susceptibilities. Finally, there is a possibility of misclassification of serotypes since they were grouped based on previous meta-serotype-specific analysis of case fatality rates [54]. However, if misclassification occurred, it is likely to bias the results towards the null, suggesting that the associations between serotypes and outcomes may be even stronger than previously reported.

## Conclusion

Based on our analysis, we have determined that Pneumococcal pneumonia significantly contributes to mortality in both adults and children. We also found that the sensitivity of blood cultures may be compromised when antibiotics have been previously administered. Therefore, it is crucial to exercise caution and prudence in the use of antibiotics.

**Figure 1.**
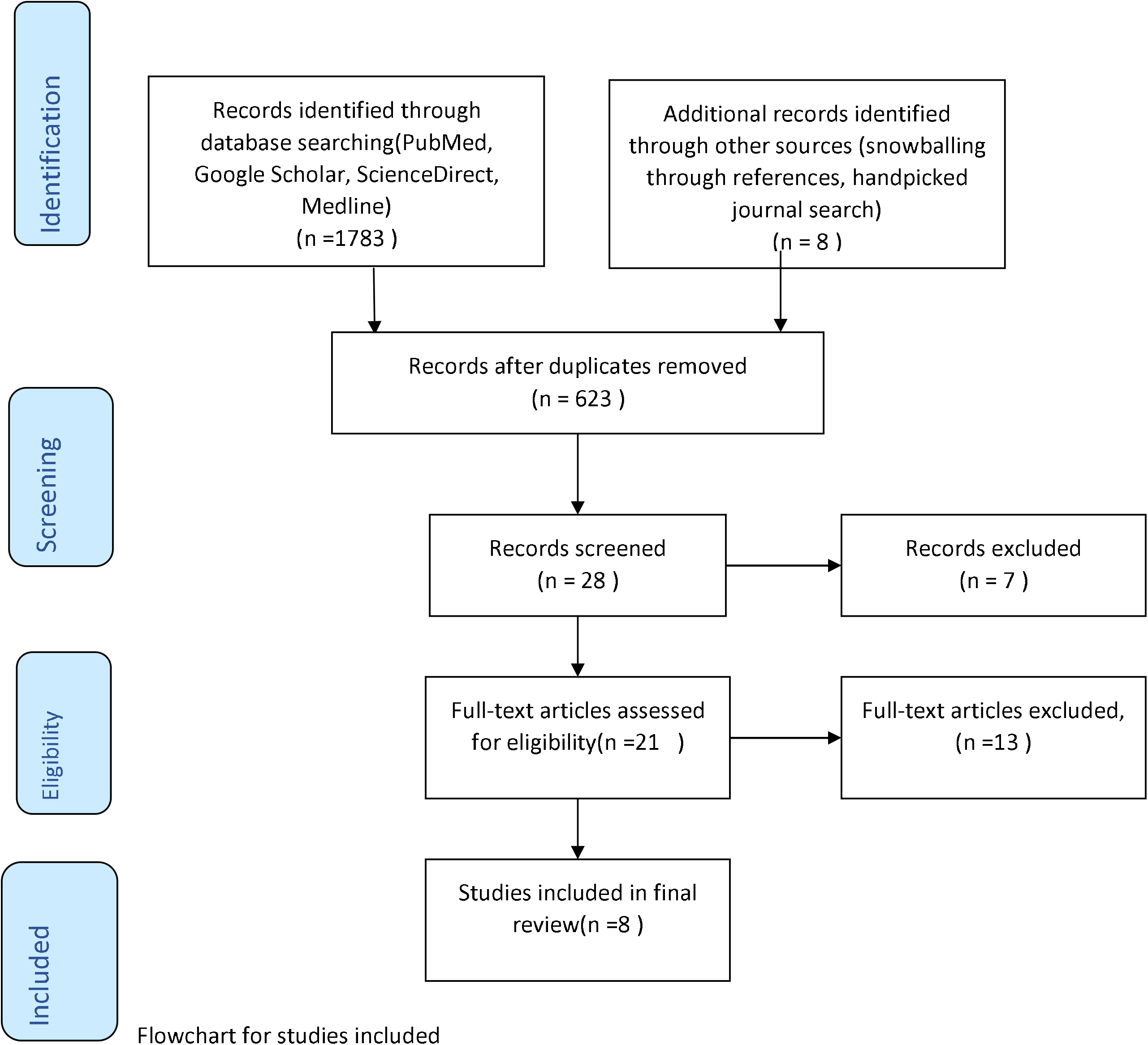
Prisma Flow Chart for study acquisition.

## Data Availability

The manuscript enclosed PRISMA table describing the studies/ research papers included and excluded in the final review.

